# Acceptability, Feasibility, and Day-to-Day Stability of a Portable Sleep Recording Device in Adolescents versus Young Adults

**DOI:** 10.1101/2025.04.13.25325346

**Authors:** Lauren S. Keller, Maya B. Fray-Witzer, Sanna Lokhandwala, Rebecca A. Hayes, Simey Chan, Peter L. Franzen, Daniel J. Buysse, Brant P. Hasler, Jessica C. Levenson, Meredith L. Wallace, Duncan B. Clark, Ronette Blake, Grace Wilson, Margaret Kuzemchak, Maria Jalbrzikowski, Adriane M. Soehner

**Affiliations:** Department of Psychiatry, University of Pittsburgh School of Medicine, Pittsburgh, PA; Department of Psychiatry and Behavioral Sciences, Boston Children’s Hospital, Boston, MA; Department of Psychiatry, Harvard Medical School, Boston, MA; Department of Pediatrics, University of Pittsburgh School of Medicine, Pittsburgh, PA; Department of Statistics, University of Pittsburgh, Pittsburgh, PA

**Author notes:** co-first author (Lauren S. Keller, Maya B. Fray-Witzer). co-senior author (Maria Jalbrzikowski, Adriane M. Soehner). **Corresponding Author:** Adriane Soehner, Ph.D. Department of Psychiatry University of Pittsburgh School of Medicine 3811 O’Hara St Pittsburgh, PA 15213. **Conflicts of Interest:** The authors have no conflicts of interest to disclose.

**Keywords:** Adolescence, Sleep Architecture, Feasibility, Acceptability

## Abstract

**Objectives:** Adolescence is marked by significant changes in sleep physiology, which provides crucial insights into brain development and health outcomes. However, sleep has been difficult to measure on a large scale due to the limitations of polysomnography. Emerging portable devices, like the Dreem3 headband (Dreem3), offer a scalable and accessible alternative, with validation studies in adults showing strong feasibility and accuracy, but validation data for adolescents remain limited. This pilot study evaluated the acceptability and feasibility of the Dreem3, and day-to-day stability of Dreem3-derived sleep estimates, in adolescents and young adults.

**Methods:** Eighty-one participants completed 3 consecutive nights of sleep recordings with the Dreem3: 32 Adolescents (9-17yrs) and 49 Young Adults (18-26yrs). We evaluated acceptability with a user experience survey and determined feasibility based on completion and quality of sleep recordings. We used intraclass correlation coefficients to assess between-night stability of sleep macro-architecture and micro-architecture estimates.

**Results:** Dreem3 was similarly acceptable for Adolescents and Young Adults (p-values>0.1) on most user experience survey outcomes except that the Young Adult group reported poorer sleep quality on Dreem3 nights relative to the Adolescent group (p<0.05). The number of recording nights (total and good quality) between-night stability of sleep architecture did not differ between groups (p-values>0.05).

**Conclusion:** Our pilot data indicates that Dreem3, is well-accepted and feasible in young people, and provide moderately stable sleep data across nights. These findings open the door to large-scale, at-home sleep studies with portable sleep recording devices in adolescents.

## Introduction

Adolescence is characterized by major developmental shifts in sleep patterns and physiology (Tarokh et al., 2016). In the second decade of life, physiological features such as slow waves or spindle activity undergo prominent changes (Colrain & Baker, 2011). Sleep physiology is a sensitive indicator of brain development (Lokhandwala & Spencer, 2022), interacts with a host of biopsychosocial factors (Afiya, 2024), and is linked to diverse physical, mental, and cognitive health outcomes (Lock et al., 2018; Luyster et al., 2012; McCarter et al., 2022). Thus, measuring sleep physiology in adolescence, and doing so in a scalable way, is vital for understanding sources of developmental variability and how aberrant health trajectories manifest. However, capturing sleep physiology at a large scale has been challenging with the current best-practice measurement approach, polysomnography (PSG). Portable, limited-montage sleep recording headbands have the potential to bridge this accessibility and scalability gap (Tobin et al., 2021), but to date there is limited feasibility and validity data for these devices in adolescents.

PSG is considered the gold-standard methodology for assessing sleep physiology (Grigg-Damberger, 2012). PSG involves simultaneous recording of electroencephalogram (EEG), electromyogram (EMG), and electrooculogram (EOG) signals during sleep. Standard PSG relies on trained staff and expensive equipment, and is labor-intensive to set up, conduct, and score, whether conducted in the lab or in-home (ambulatory). These features make it impractical to gather multi-night, home-based PSG recordings on a large scale (Rundo & Downey, 2019) The ability to obtain multi-night home-based sleep recordings would provide a more ecologically valid picture of sleep physiology. Among adolescents, it is becoming increasingly apparent that multi-night recordings are needed to obtain stable and valid sleep EEG estimates. Although there are some heritable and trait-like aspects of the sleep EEG (Rusterholz et al., 2018; Tarokh et al., 2011), sleep loss, sleep fragmentation, or sleep timing shifts – common occurrences in adolescents (Crowley et al., 2018) – can introduce variability in sleep stage duration and sleep micro-architecture (Ong et al., 2019), reducing the reliability of sleep endpoints drawn from a single-night recording.

Emerging data indicates that portable sleep recording devices offer an accurate, cost-effective, and accessible alternative to PSG. While the designs of portable sleep recording devices (e.g., Dreem3, CGX, Zmachine, SleepProfiler, Muse S) varies, most involve a limited EEG montage headband without EOG and EMG (Alnosh et al., 2023). These devices are simple enough for a non-expert to use, and some come with accompanying mobile, cloud-based, or computer applications which stage sleep automatically. Finally, most portable sleep recording devices can be used at home, across multiple nights. Overall, these features reduce participant burden, better capture habitual sleep, and increase the accessibility of sleep physiology recordings. In validation studies conducted in adults, many of these devices report strong feasibility and acceptability, and show high agreement between automated sleep staging algorithms and traditional visual sleep staging by expert technicians (Chinoy et al., 2022; Krigolson et al., 2017; Levendowski et al., 2017). However, only one study to date has demonstrated initial feasibility in an adolescent population (Lunsford-Avery et al., 2020). This study did not evaluate aspects of sleep EEG signal quality or between-night stability of sleep outcomes (Lunsford-Avery et al., 2020).

Our goal was to evaluate the acceptability, feasibility, and between-night stability of Dreem3 headband recordings (Dreem3) in adolescents (9-17yrs) and young adults (18-26yrs). In small validation studies conducted in adults (N=60), the Dreem3 automated sleep staging algorithm showed high agreement with expert scorers (Ong et al., 2024) and strong coherence with simultaneously measured PSG (Arnal et al., 2020). Acceptability and feasibility of the Dreem3 has also been demonstrated in multiple adult cohorts (Arnal et al., 2020; González et al., 2024; Ong et al., 2024). We hypothesized that the Dreem3 would have high acceptability ratings as well as limited impact on sleep and next day disturbances in alertness, mood, and concentration. Similarly, we hypothesized that Dreem3 recordings would show similar feasibility in adolescents and young adults, that we would be able to obtain a high proportion of recording nights and high-quality data in both groups. Lastly, between-night stability of sleep staging and quantitative sleep EEG outcomes were evaluated. We expected to find that the sleep outcomes would be more stable from between-night in young adults than in adolescents, highlighting the importance of multi-night recordings to obtain stable sleep estimates in youth.

## Methods

### Participants

This sample included healthy participants without bipolar spectrum disorder, psychosis spectrum disorder, or neurodevelopmental conditions. Participants were permitted to be on a stable psychotropic medication regimen **(Figure 1)**.

**Figure 1.**
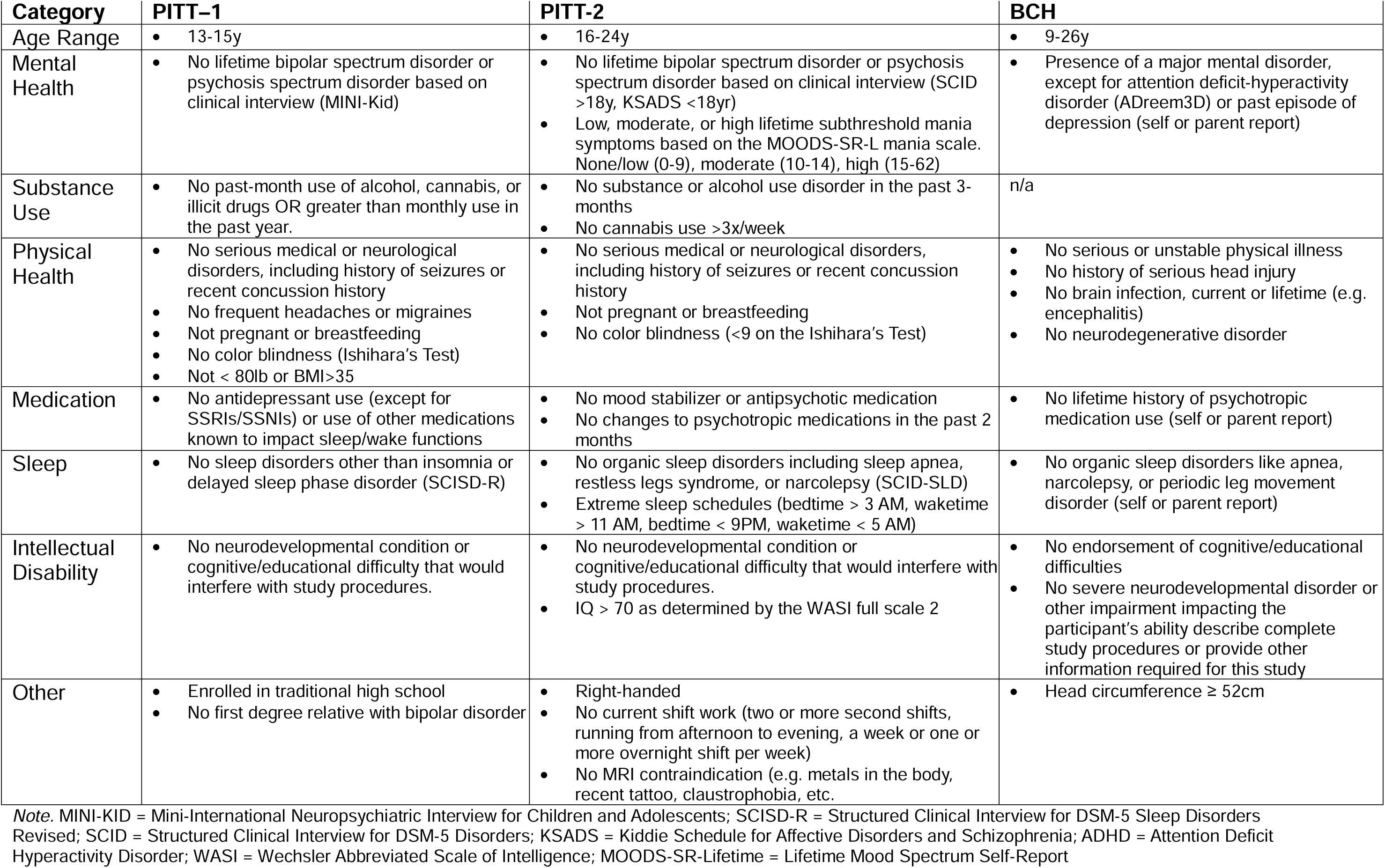
Eligibility criteria across the three study samples. All participants had to be fluent in English and willing/able to engage in all research procedures.

#### University of Pittsburgh-Study 1 (PITT-1)

Twenty-three healthy adolescents (13-16 years-old) were enrolled in a Dreem3 pilot study at the University of Pittsburgh. Participants were recruited from a larger parent protocol (P50DA046346) focused on understanding relationships between sleep-circadian rhythms and substance use risk. Participants from the parent study were recruited from a local participant registry, community advertisements, newsletters, and social media posts.

#### University of Pittsburgh-Study 2 (PITT-2)

Forty-five adolescents and young adults (16-24 years-old) were enrolled in a second Dreem3 pilot study at the University of Pittsburgh. Participants were recruited from a larger parent protocol focused on understanding sleep-circadian rhythms and mood disorder risk (R01MH124828). This study used recruitment methods similar to PITT-1.

#### Boston Children’s Hospital (BCH)

Eighteen healthy individuals (9-26 years-old) were recruited for a Dreem3 pilot study through flyers, previous study participation, the Precision Link and Biobank (PLB, (Bourgeois et al., 2017), and online posts.

#### Analytic Sample

Across the three combined studies, 81 participants were considered for the main analyses: 32 Adolescents (9-17yrs) and 49 Young Adults (18-26yrs). In PITT-1 (N=23), two participants were excluded from all analyses due to missing Dreem and UX data. In PITT-2 (N=45), Beacon’s cloud service down-time prevented one participant from completing the protocol and two participants were excluded due to missing Dreem Data. We included all participants from the BCH sample (N=18). We had a final total analytic sample of N=81.

User experience survey analyses evaluating acceptability outcomes were conducted in 66 participants; fifteen participants (5 PITT-1, 5 PITT-2, 5 BCH) were excluded due to missing survey data. Feasibility outcomes were assessed in all 81 participants. We evaluated between-night stability of sleep outcomes in a subset of 72 participants (N=19 PITT-1, N=36 PITT-2, N=17 BCH) who had at least two nights of good quality recordings, based on quality control procedures reported below.

### Key Measures

#### Dreem3 Headband (Dreem3)

The Dreem3 is a wireless sleep EEG recording device developed by Dreem, Inc. (Paris, FR) and owned by Beacon Biosignals, Inc. (Boston, MA). The Dreem3 is plastic and foam with an adjustable elastic band and Velcro clasp, accommodating head sizes of at least 50 cm. It includes two frontal EEG sensors (F7, F8), one ground sensor (Fp2), and two occipital sensors (O1, O2) to record cortical electrical activity. Additionally, the Dreem3 can measure bodily movement, head positioning, and respiration via a built-in accelerometer, and heart rate via a pulse oximeter. After collection, sleep EEG data is sent via Bluetooth from the headband to the Alfin app installed on the participants’ mobile device, and then uploaded for storage and analysis on Beacon’s cloud-based website. This device is equipped to record up to 200 hours of sleep data.

#### User Experience Survey

The self-report user experience survey consisted of 23 questions (**Figure S1**) designed to evaluate aspects of the Dreem3 and Alfin app experience. Ease of use (Dreem3 and Alfin app) and comfort (Dreem3) were rated from 1 (very poor) to 7 (very good). Next-day impacts of wearing the Dreem3 (mood, alertness, irritability, concentration) and same-night sleep quality were rated on a Likert-scale ranging from -2 (much worse) to 2 (much better). Acceptability of the 3-night tracking duration was rated on a scale of 1 (much too short) to 7 (much too long). Participants also rated the likelihood of recommending the Dreem3 to a friend on a scale of 1 (strongly not recommend) to 7 (strongly recommend) and overall satisfaction on a scale from 1 (very dissatisfied) to 7 (very satisfied).

### Procedures

For PITT studies, all study procedures were approved by the Institutional Review Board at the University of Pittsburgh, Pittsburgh Campus prior to study start and all in-lab study procedures took place at the University of Pittsburgh Medical Center Western Psychiatric Hospital. For BCH, all study procedures were approved by the Boston Children’s Hospital Institutional Review Board.

After consent (18+) or assent (<18yrs), participants completed study-specific eligibility evaluation procedures and a survey battery. Participants who opted into the Dreem3 studies were taught how to use the Dreem3 and completed daily electronic sleep diaries. In PITT-1 and BCH, participants completed 3 consecutive nights of overnight Dreem3 sleep monitoring at self-selected habitual sleep times at home. In PITT-2, participants completed four consecutive nights of Dreem3 monitoring: one in lab (to instruct participants on wearing the device), two at-home, and one final night of concurrent Dreem3-PSG recording. We used the first 3 of these nights in the present analyses. After completing the final night of Dreem3 sleep monitoring, all participants completed the user experience questionnaire. All participants were compensated as part of the study procedures.

### Sleep Data Processing

#### Sleep Staging

American Academy of Sleep Medicine (AASM) sleep stage classification (Iber, 2007) was performed using the Dreem3 automated algorithm, which has been validated against PSG and manual sleep staging by expert technicians in healthy adult participants (Arnal et al., 2020). The Dreem3 sleep staging algorithm uses a combination of features derived from EEG, pulse oximeter, and accelerometer signals to determine the probability that each epoch belongs to each sleep stage. To predict the sleep stage for a given epoch, algorithms incorporate features extracted from the current and past 30 epochs. In a validation study of adults, the sleep staging algorithm correctly classified sleep stages at a level equivalent to manual sleep scoring by experts (83.8%) (Arnal et al., 2020).

#### Quantitative Sleep EEG Analyses

##### EEG preprocessing

Sleep EEG data were processed using the open-source package *Luna* (http://zzz.bwh.harvard.edu/luna/). Dreem3 sleep EEG was sampled at 250Hz and band-pass filtered (0.3Hz, 35Hz) in the original records. Analyses focused on the F7-O2 and F8-O1 channels. In *Luna,* all EEG signals were first converted to uV. Within NREM (N2, N3) and REM epochs (based on Dreem3 automated staging), we identified and removed epochs with artifact and/or signal outliers based on the following criteria: a) delta power greater than 2.5 times the local average or beta power more than 2.0 times the local average, identified using Welch’s method, b) maximum amplitudes over 200 uV for more than 5% of the epoch, c) flat or clipped signals for more than 5% of the epoch, and d) signals 3.5 standard deviations from the mean for any of the three Hjorth parameters (i.e., activity, mobility, and complexity; Hjorth, 1970). Outlier removal based on Hjorth parameters was performed twice for each record, based on prior work (Kozhemiako et al., 2024). Channels were excluded if >50% of the epochs contained identified artifacts. In addition, records were excluded if there was <180 min of total sleep time (TST). After artifact rejection, we also used *Luna* to perform power spectral analysis, spindle detection, and slow oscillation (SO) detection.

##### Spectral power estimation

Welch’s method was used to estimate spectral power for NREM and REM separately. For each 30-second epoch, we applied a Fast Fourier Transform using 4-second segments (0.25 spectral resolution) windowing with a Tukey taper (50%); consecutive segments overlapped by 50% (2 seconds). Spectral power was summarized in traditional frequency bands: delta [1-4Hz], theta [4-8 Hz], alpha [8-12 Hz], sigma [12-15 Hz], and beta [15-30 Hz]. For each channel (F8-O1, F7-O2), we computed average power in each band for each epoch, then across NREM (N2, N3) and REM epochs separately. Relative power for each band was computed with respect to the total absolute power.

##### Spindle detection

Consistent with other reports (Kozhemiako et al., 2024; Purcell et al., 2017; Warby et al., 2014) we detected spindles in the “slow” (11 Hz) and “fast” (15 Hz) frequencies. We computed spindle density (number of spindles per minute), amplitude (based on maximum peak-to-peak amplitude), duration (seconds), mean integrated spindle activity per spindle (average amplitude and duration of an individual’s spindles), spindle chirp (mean rate of frequency within a spindle), Fast Fourier Transform, and mean spindle frequency.

##### Slow oscillation (SO) detection

SOs were identified based on the EEG signals band-pass filtered between 0.5 and 4 Hz using default *Luna* settings (as described in (Djonlagic et al., 2021; Kozhemiako et al., 2024). The following temporal criteria were used to define SOs: 1) a consecutive zero-crossing leading to negative peak was between 0.5 and 1.5 seconds; and 2) a zero-crossing leading to positive peak was not longer than 1 seconds. Two approaches were used to measure SO amplitude: 1) an adaptive/relative threshold required that negative peak and peak-to-peak amplitudes be greater than twice the mean (for that individual/channel) and 2) an absolute threshold required a negative peak amplitude larger than -40 mV, and peak-to-peak amplitude larger than 75 mV. SO density (count per minute), mean amplitude of the negative peak, peak-to-peak amplitude, duration, and the upward slope of negative peak were estimated for each channel.

#### Sleep Recording Quality Assessment

For sleep staging, a good quality sleep staging for Dreem3 was defined as at least 180 min of recording time (to obtain approx. two NREM-REM cycles). The *Luna* pipeline identified 30-second epochs with and without artifact (described above). For quantitative outcomes, data from each EEG derivation were rated as good (>50% sleep epochs without artifact), fair (30-49% of sleep epochs without artifact), and poor quality (<30% of sleep epochs without artifact).

### Data Analysis

#### Data harmonization

Data were harmonized across the three cohorts and split into two age groups: Adolescent (9-17yrs, n=32) and Young Adult (18-26yrs; n=49). Age group differences in participant characteristics are described in **Table 2** using chi-square test (or Fisher’s exact test) and t-tests.

#### Acceptability

We used the UX survey to measure Dreem3 acceptability. We used t-tests to conduct age group comparisons between Adolescents vs Young Adults. Acceptability outcomes included: user experience with the app and device (ease of device and app setup, comfort, ease of sleep, device application, overall satisfaction), as well as effects on sleep and next-day functioning (sleep quality, mood, alertness, daytime sleepiness, irritability, concentration). We also explored Pearson correlations between age and UX acceptability outcomes.

#### Feasibility

Three metrics were used to evaluate Dreem3 feasibility: the proportion of completed recording nights (attempted recording nights / total recording nights in protocol), the proportion of nights with good quality automated sleep staging data, and the proportion of nights with good quality quantitative EEG data, as defined in “Sleep Recording Quality Assessment” above. T-tests compared feasibility between the two age groups. We also explored associations between age and feasibility outcomes using Spearman’s rank correlations.

#### Between-night stability of sleep estimates

Intraclass Correlation Coefficients (ICCs) were used to test within-subject between-night stability of Dreem3 sleep staging, and quantitative sleep EEG outcomes. We conducted mixed-effects ICC models on good quality recordings, adjusting for the percentage of artifact-free epochs in each record. ICCs were classified as poor (<0.50), moderate (0.50-0.75), good (0.75-0.90), or excellent (>0.90) based on established guidelines (Koo & Li, 2016). ICCs ≥ 0.60 indicate reliable stability. Group differences (Adolescent vs Young Adult) were compared using bootstrapped 95% confidence intervals (500 iterations); non-overlapping confidence intervals were considered significantly different.

#### First-Night effect

Sleep quality can be worse on the first night of in-lab PSG recordings (Byun et al., 2019). To evaluate group differences in the ‘first night’ effect for the Dreem3, linear mixed effects models evaluated whether there was a group (Adolescent, Young Adult) by night interaction effect on sleep efficiency as an index of sleep quality, adjusting for study (PITT-1, PITT-2, BCH).

## Results

### Participant Characteristics

Participant characteristics by age group (Adolescent vs. Young Adult) are described in **Table 1** and by study in **Table S1.** Adolescent and Adult groups did not differ in sex at birth or ethnicity. There were age group differences in race, sleep tracking on weekdays versus weekends, and psychotropic medication use (p-values<0.05). Asian race was more common in the Young Adult group relative to the Adolescent Group. Sleep tracking on weekdays was higher in the Adolescent group whereas weekend sleep tracking was higher in the Young Adult group. Psychotropic medication use was more common in the Young Adult group than the Adolescent group.

**Table 1.** Participant demographic and clinical characteristics by age group, N=81.

| **Variable** | **Adolescents**<br>**(9-17y; n=32)** | **Young Adults**<br>**(18-26y; n=49)** | **Group Comparison** |  |
| --- | --- | --- | --- | --- |
|  | *Mean(SD) or N(%)* | *Mean(SD) or N(%)* | *Statistic* | *p-value* |
| Age (yr) | 14.26 (1.97) | 22.55 (1.73) | t(61)=-19.40 | **p<0.001** |
| Female at Birth | 18 (56.25) | 27 (55.01) | X<sup>2</sup>(1)=0.00 | p=1 |
| Hispanic/Latino | 4 (12.50) | 1 (2.04) | X<sup>2</sup>(1)=2.07 | p=0.15 |
| Race |  |  | X<sup>2</sup>(3)=13.92 | **p=0.003** |
| Asian | 0 (0.00) | 16 (32.65) | -- | -- |
| Black | 3 (9.38) | 4 (8.16) | -- | -- |
| Multiple Races | 5 (15.63) | 2 (4.08) | -- | -- |
| White | 24 (75.00) | 27 (55.01) | -- | -- |
| Sleep Tracking |  |  | X<sup>2</sup>(1)= 13.52 | **p<0.001** |
| Weekends | 0.66 (0.87) | 1.45 (0.79) | -- | -- |
| Weekdays | 2.50 (1.27) | 1.86 (0.58) | -- | -- |
| Psychotropic Medication | 0 (0.00) | 8 (16.32) | X<sup>2</sup>(1)=4.10 | **p=0.04** |
| Study |  |  |  |  |
| PITT-Study 1 | 21 (65.63) | 0 (0.00) | -- | -- |
| PITT-Study 2 | 3 (9.38) | 39 (79.59) | -- | -- |
| BCH | 8 (25.00) | 10 (20.41) | -- | -- |

*Note:* yr = year; PITT = University of Pittsburgh; BCH = Boston Children's Hospital; SD = standard deviation; N = number

### Age Group Differences in Acceptability

User experience outcomes are described by group in **Table 2** and plotted in **Figure 2**. The two groups did not significantly differ in ratings of their experience with the app and device (ease of device and app setup, comfort, ease of sleep, device application, overall satisfaction) and the impact on next-day mood, alertness, sleepiness, irritability, or concentration (all p-values>0.10). The Young Adult group rated their sleep quality on Dreem3 nights as worse than the Adolescent group (p=0.02). In exploratory analyses, age was negatively correlated with self-reported sleep quality (r=-0.28, p=0.02), but there were no other statistically significant correlations between age and Dreem3 acceptability (**Table S2)**.

**Figure 2.**
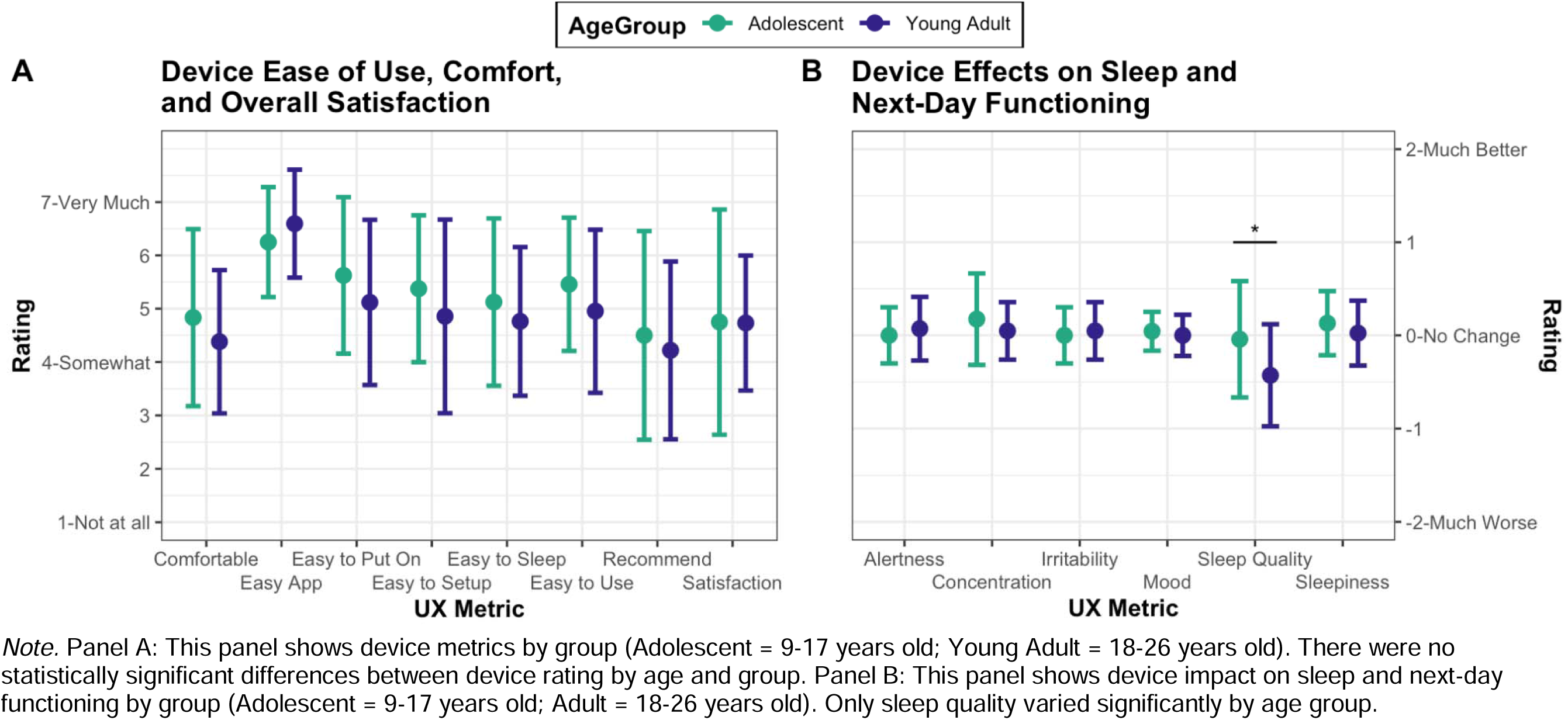
UX Constructs by Age Group

**Table 2.** User Experience (UX) survey acceptability data by age group, N=66.

| UX Variable | Adolescents<br>(9-17y; n=24) |  | Young Adults<br>(18-26y; n=42) |  | Statistic |  |
| --- | --- | --- | --- | --- | --- | --- |
|  | Mean | SD | Mean | SD | t-statistic | p-value |
| Mobile application was easy to use (Easy App)<sup>a</sup> | 6.25 | 1.03 | 6.60 | 1.01 | -1.32 | 0.19 |
| Dreem3 was easy to set up in the mobile application (Easy to Set Up)<sup>a</sup> | 5.38 | 1.38 | 4.86 | 1.82 | 1.30 | 0.20 |
| Dreem3 was easy to sleep with on the first night (Easy to Sleep)<sup>a</sup> | 5.13 | 1.57 | 4.76 | 1.39 | 0.94 | 0.35 |
| Dreem3 was comfortable (Comfortable)<sup>a</sup> | 4.83 | 1.66 | 4.38 | 1.34 | 1.14 | 0.26 |
| Dreem3 was easy to put on (Easy to Put On)<sup>a</sup> | 5.63 | 1.47 | 5.12 | 1.55 | 1.32 | 0.19 |
| Dreem3 was easy to use (Easy to Use)<sup>a</sup> | 5.46 | 1.25 | 4.95 | 1.53 | 1.46 | 0.15 |
| Would recommend Dreem3 to a friend (Recommend)<sup>a</sup> | 4.50 | 1.96 | 4.22 | 1.67 | 0.59 | 0.56 |
| Satisfaction wearing Dreem3 (Satisfaction)<sup>a</sup> | 4.75 | 2.11 | 4.73 | 1.27 | 0.04 | 0.97 |
| Sleep Quality<sup>b</sup> | -0.04 | 0.62 | -0.43 | 0.55 | 2.53 | **0.02** |
| Mood<sup>b</sup> | 0.04 | 0.21 | 0.00 | 0.22 | 0.79 | 0.44 |
| Alertness<sup>b</sup> | 0.00 | 0.30 | 0.07 | 0.34 | -0.87 | 0.39 |
| Sleepiness<sup>b</sup> | 0.13 | 0.34 | 0.02 | 0.35 | 1.19 | 0.24 |
| Irritability<sup>b</sup> | 0.00 | 0.30 | 0.05 | 0.31 | -0.60 | 0.55 |
| Concentration<sup>b</sup> | 0.17 | 0.49 | 0.05 | 0.31 | 1.12 | 0.27 |

Note. For the first eight items, scales ranged from 1 - 7. For the last six items, scales ranged from -2 – 2

Given that race, weekday vs. weekend sleep tracking days, and psychotropic medication use differed between age groups, exploratory ANCOVA models evaluated whether covarying for these variables impacted acceptability outcomes (see **Table S4).** With these variables included as covariates, the difference in sleep quality ratings between Adolescents and Young Adults fell just below significance (p=0.06), and a group difference emerged in the ease of mobile application use (p=0.03), with Adolescents rating the app as easier to use than Young Adults. Age group differences were not statistically significant for any other metrics.

### Age Group Differences in Feasibility

There was no significant difference in completed recording nights (nights of attempted Dreem3 recording vs total recording Dreem3 nights in protocol) between age groups (Adolescent 2.75±0.10 nights vs Young Adult 2.78±0.08 nights, p=0.85). Additionally, the groups did not significantly differ with respect to number of nights with good quality sleep staging (Adolescent 2.59±0.71 nights vs Young Adult 2.80±0.45 nights, p=1.00) nor did they significantly differ with respect to the number of nights of good quality quantitative EEG data (F7 – Adolescent 2.44±0.76 nights vs Young Adult 2.72±0.58 nights, p=0.56; F8 – Adolescent 2.44±0.76 nights vs Young Adult 2.59±0.69 nights, p=0.42; **Fig 3**.). The correlation between age and number of recorded nights completed was small and not significant (*rho*=0.06, p=0.59).

**Figure 3.**
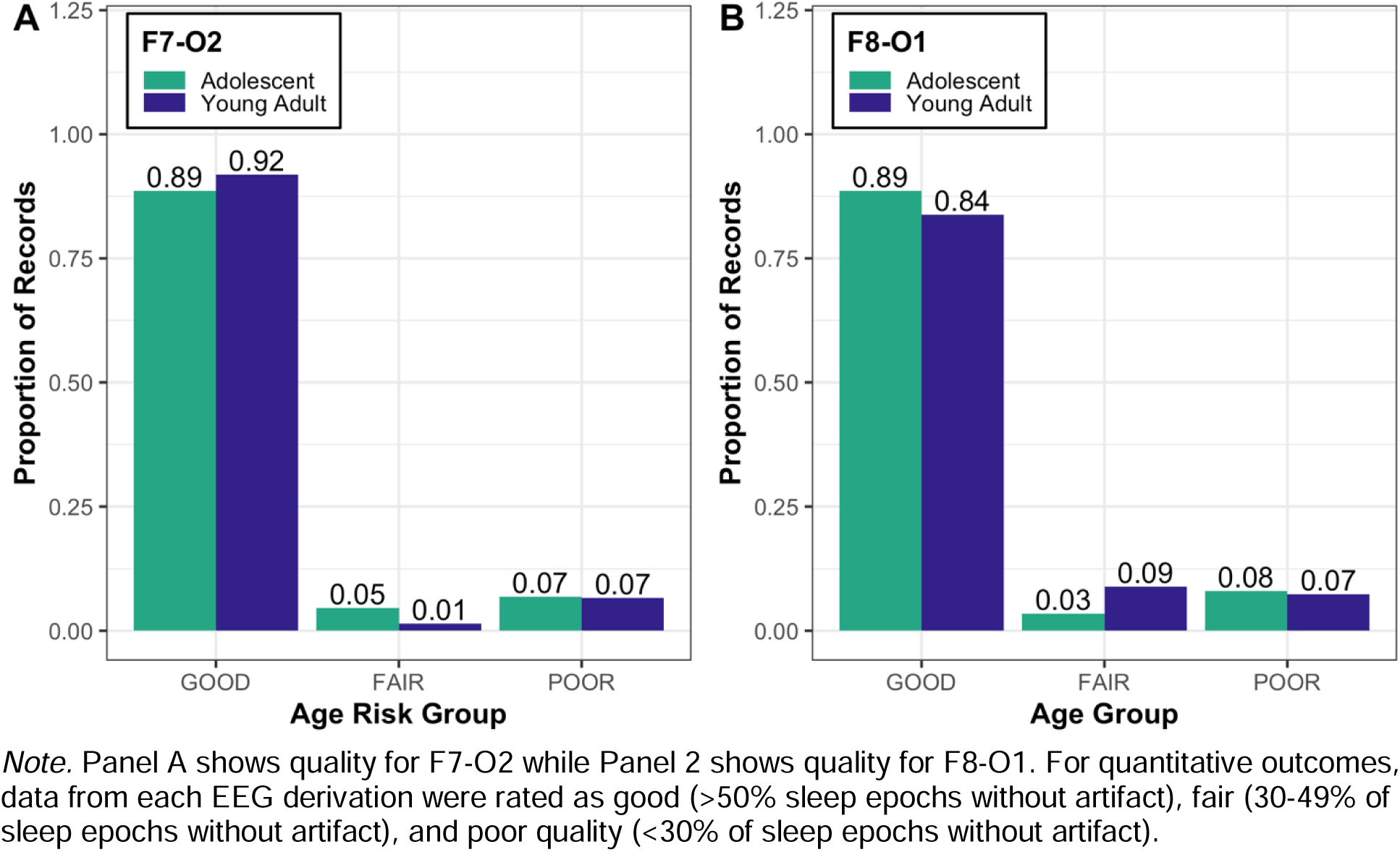
Quantitative EEG outcome quality for F7-O2 and F8-O1 derivations, by age group

### Age Group Differences in Between-Night Stability

For participants with good quality F7-O2 data across at least two nights (n=72), group differences for the between-night stability of Dreem3-derived sleep estimates are described in **Table 3** and plotted in **Figure 4. Table S3** reports ICCs for F8-O1 which are similar to F7-O2. The ICC estimates did not differ between age groups for any sleep estimates (all bootstrapped 95% confidence intervals overlapped). Most sleep variables had moderate between-night stability (ICCs 0.50-0.75). Overall, spindle characteristics showed the strongest between-night stability across groups (0.18-0.79). A few variables demonstrated poor stability (ICCs < 0.50), such as REM%, TST Hours, and TWT Hours. Between-night stability of sleep estimates by age group for participants with at least 3 nights of good quality sleep data (F7-O2 and F8-O1 derivations) can be found in **Figure S4.**

**Figure 4.**
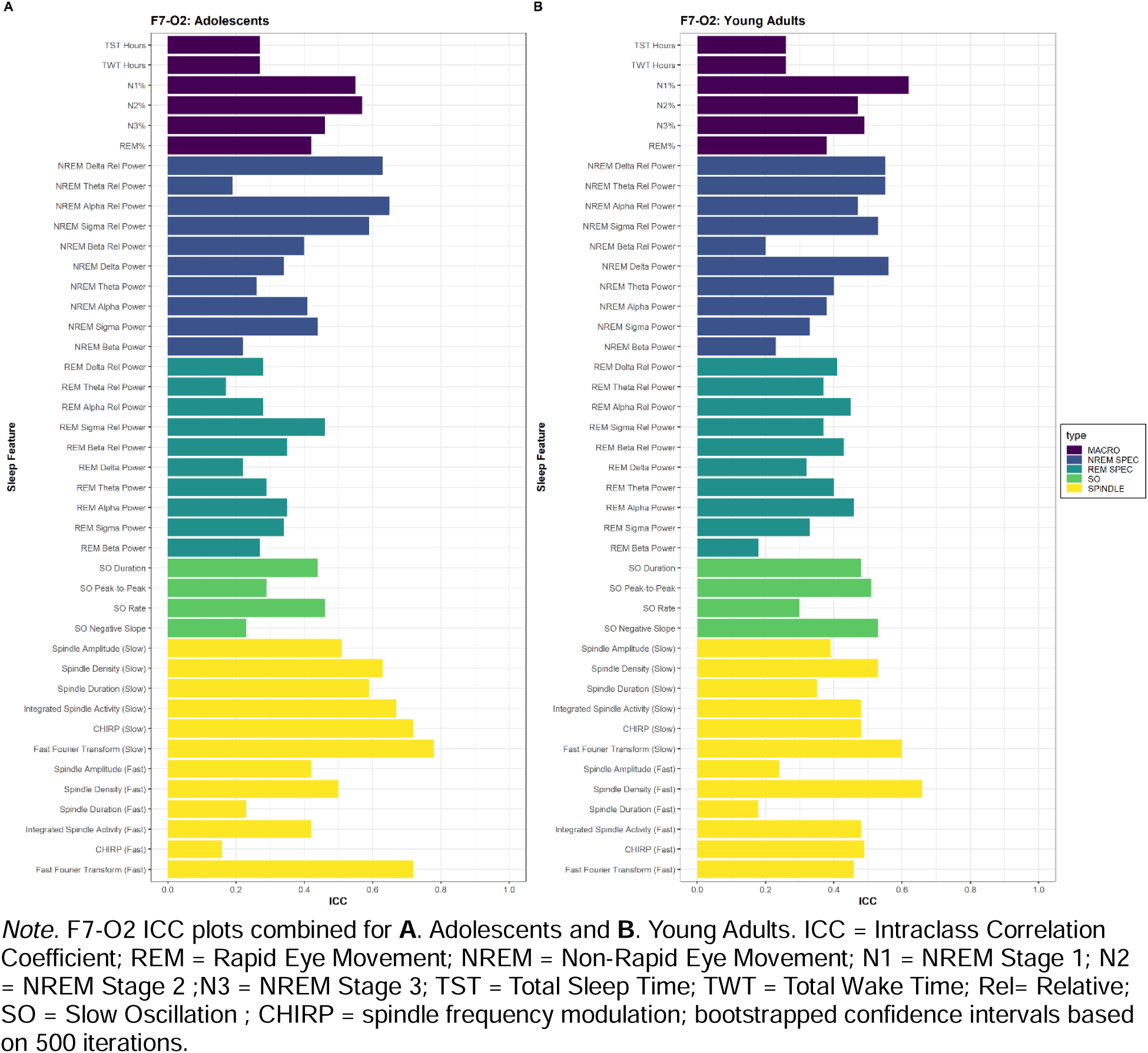
Between-night stability (ICC) of sleep estimates by age group for participants with at least 2 nights of good quality sleep data (F7-O2 derivation)

**Table 3:**
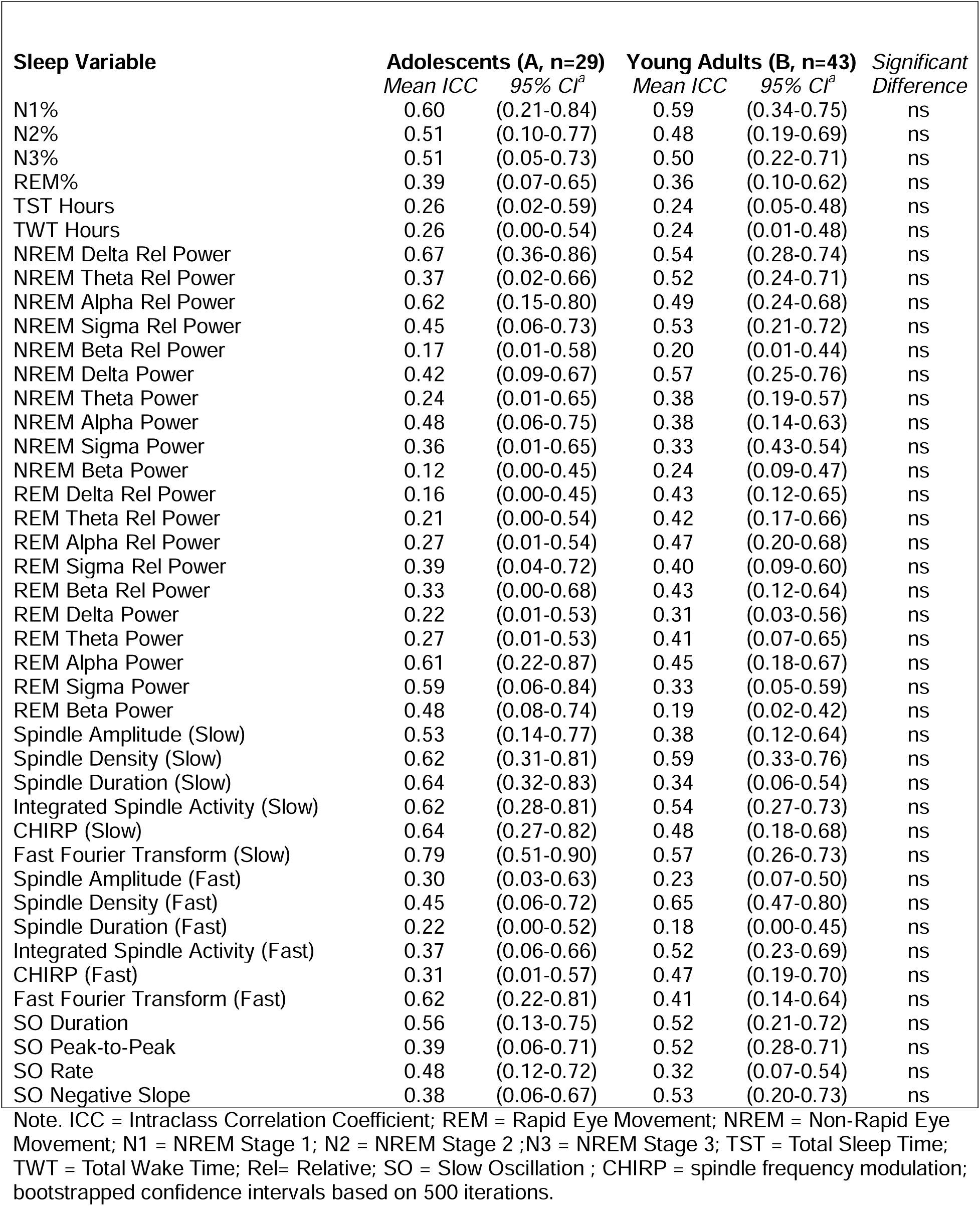
Between-night stability (ICC) of sleep estimates by age group for participants with at least 2 nights of good quality sleep data for F7-O2 (n=72).

| Sleep Variable | Adolescents (A, n=29) |  | Young Adults (B, n=43) |  | Significant Difference |
| --- | --- | --- | --- | --- | --- |
|  | Mean ICC | 95% CI<sup>a</sup> | Mean ICC | 95% CI<sup>a</sup> |  |
| N1% | 0.60 | (0.21-0.84) | 0.59 | (0.34-0.75) | ns |
| N2% | 0.51 | (0.10-0.77) | 0.48 | (0.19-0.69) | ns |
| N3% | 0.51 | (0.05-0.73) | 0.50 | (0.22-0.71) | ns |
| REM% | 0.39 | (0.07-0.65) | 0.36 | (0.10-0.62) | ns |
| TST Hours | 0.26 | (0.02-0.59) | 0.24 | (0.05-0.48) | ns |
| TWT Hours | 0.26 | (0.00-0.54) | 0.24 | (0.01-0.48) | ns |
| NREM Delta Rel Power | 0.67 | (0.36-0.86) | 0.54 | (0.28-0.74) | ns |
| NREM Theta Rel Power | 0.37 | (0.02-0.66) | 0.52 | (0.24-0.71) | ns |
| NREM Alpha Rel Power | 0.62 | (0.15-0.80) | 0.49 | (0.24-0.68) | ns |
| NREM Sigma Rel Power | 0.45 | (0.06-0.73) | 0.53 | (0.21-0.72) | ns |
| NREM Beta Rel Power | 0.17 | (0.01-0.58) | 0.20 | (0.01-0.44) | ns |
| NREM Delta Power | 0.42 | (0.09-0.67) | 0.57 | (0.25-0.76) | ns |
| NREM Theta Power | 0.24 | (0.01-0.65) | 0.38 | (0.19-0.57) | ns |
| NREM Alpha Power | 0.48 | (0.06-0.75) | 0.38 | (0.14-0.63) | ns |
| NREM Sigma Power | 0.36 | (0.01-0.65) | 0.33 | (0.43-0.54) | ns |
| NREM Beta Power | 0.12 | (0.00-0.45) | 0.24 | (0.09-0.47) | ns |
| REM Delta Rel Power | 0.16 | (0.00-0.45) | 0.43 | (0.12-0.65) | ns |
| REM Theta Rel Power | 0.21 | (0.00-0.54) | 0.42 | (0.17-0.66) | ns |
| REM Alpha Rel Power | 0.27 | (0.01-0.54) | 0.47 | (0.20-0.68) | ns |
| REM Sigma Rel Power | 0.39 | (0.04-0.72) | 0.40 | (0.09-0.60) | ns |
| REM Beta Rel Power | 0.33 | (0.00-0.68) | 0.43 | (0.12-0.64) | ns |
| REM Delta Power | 0.22 | (0.01-0.53) | 0.31 | (0.03-0.56) | ns |
| REM Theta Power | 0.27 | (0.01-0.53) | 0.41 | (0.07-0.65) | ns |
| REM Alpha Power | 0.61 | (0.22-0.87) | 0.45 | (0.18-0.67) | ns |
| REM Sigma Power | 0.59 | (0.06-0.84) | 0.33 | (0.05-0.59) | ns |
| REM Beta Power | 0.48 | (0.08-0.74) | 0.19 | (0.02-0.42) | ns |
| Spindle Amplitude (Slow) | 0.53 | (0.14-0.77) | 0.38 | (0.12-0.64) | ns |
| Spindle Density (Slow) | 0.62 | (0.31-0.81) | 0.59 | (0.33-0.76) | ns |
| Spindle Duration (Slow) | 0.64 | (0.32-0.83) | 0.34 | (0.06-0.54) | ns |
| Integrated Spindle Activity (Slow) | 0.62 | (0.28-0.81) | 0.54 | (0.27-0.73) | ns |
| CHIRP (Slow) | 0.64 | (0.27-0.82) | 0.48 | (0.18-0.68) | ns |
| Fast Fourier Transform (Slow) | 0.79 | (0.51-0.90) | 0.57 | (0.26-0.73) | ns |
| Spindle Amplitude (Fast) | 0.30 | (0.03-0.63) | 0.23 | (0.07-0.50) | ns |
| Spindle Density (Fast) | 0.45 | (0.06-0.72) | 0.65 | (0.47-0.80) | ns |
| Spindle Duration (Fast) | 0.22 | (0.00-0.52) | 0.18 | (0.00-0.45) | ns |
| Integrated Spindle Activity (Fast) | 0.37 | (0.06-0.66) | 0.52 | (0.23-0.69) | ns |
| CHIRP (Fast) | 0.31 | (0.01-0.57) | 0.47 | (0.19-0.70) | ns |
| Fast Fourier Transform (Fast) | 0.62 | (0.22-0.81) | 0.41 | (0.14-0.64) | ns |
| SO Duration | 0.56 | (0.13-0.75) | 0.52 | (0.21-0.72) | ns |
| SO Peak-to-Peak | 0.39 | (0.06-0.71) | 0.52 | (0.28-0.71) | ns |
| SO Rate | 0.48 | (0.12-0.72) | 0.32 | (0.07-0.54) | ns |
| SO Negative Slope | 0.38 | (0.06-0.67) | 0.53 | (0.20-0.73) | ns |

Note. ICC = Intraclass Correlation Coefficient; REM = Rapid Eye Movement; NREM = Non-Rapid Eye Movement; N1 = NREM Stage 1; N2 = NREM Stage 2 ;N3 = NREM Stage 3; TST = Total Sleep Time; TWT = Total Wake Time; Rel= Relative; SO = Slow Oscillation ; CHIRP = spindle frequency modulation; bootstrapped confidence intervals based on 500 iterations.

### “First Night” Effect

We found no differences in sleep efficiency between recording nights or age groups (**Table 4**); a “first night” effect was not observed.

**Table 4:** Mixed-effect model testing differences in sleep efficiency between nights by age group to evaluate the “first-night” effect, adjusting for sample (PITT-2, PITT-2, BCH).

| **Variable** | ***F*-statistic** | ***p*-value** |
| --- | --- | --- |
| Sample | 1.318 | 0.274 |
| Age Group | 0.517 | 0.475 |
| Night | 0.015 | 0.985 |
| Age Group x Night | 2.081 | 0.129 |

*Note.* SE = standard error.

## Discussion

We assessed the Dreem3’s acceptability, feasibility, and between-night-stability in adolescents versus young adults over 3 consecutive nights. As hypothesized, both groups reported above-average satisfaction. However, young adults reported poorer sleep quality on Dreem3 nights. The Dreem3 demonstrated high feasibility, with both groups completing at least two of the three recording nights on average. Contrary to our hypothesis, there were no age differences in between-night stability. Most sleep variables showed moderate stability across age groups.

We found Dreem3 usage to be acceptable to both age groups. UX scores averaged above “somewhat” easy to use, likely to recommend, etc. The main between-group difference was that the young adult group reported poorer sleep quality. This may be explained by young adults’ increased likelihood to experience poor sleep quality in general (Afandi et al., 2013; Lueke & Assar, 2024; Lund et al., 2010). The college lifestyle, changes in parental supervision, and greater work responsibilities all provide a pre-disposition for stress-induced sleep difficulties. It is also possible that subjective sleep quality differed between the two groups due to data collection differences between studies. Further, it is possible that a “first night effect” exists in young adults as with PSG studies (Byun et al., 2019). An adaptation night might be advisable for portable sleep EEG recordings but, of note, we found no differences in sleep efficiency between nights or age groups in an exploratory analysis. Future studies should examine such effects for portable sleep EEG devices and should include detailed questions about device impact on sleep quality.

The feasibility of the Dreem3 headband was promising. Both groups tended to complete most recording nights (averaging >2 nights out of 3). Other studies of portable sleep EEG devices either evaluated only one night of recording (Arnal et al., 2020; González et al., 2024; Ong et al., 2024) or studied more nights (usually seven) but did not formally evaluate feasibility (Chinoy et al., 2022). One study evaluating the feasibility of a different portable sleep EEG device over multiple nights reported similar findings to those observed here, with a high rate (94.2%) of completion across seven nights of tracking in adolescents (Lunsford-Avery et al., 2020). In addition to high completion rates, most completed recordings in our study also had good quality sleep staging and quantitative EEG data. Previous studies that evaluated quantitative portable EEG data quality in adults are in line with our current findings, indicating comparability to devices such as the Muse (Arnal et al., 2020; Krigolson et al., 2017). Our results demonstrate that Dreem3 recording completion rates and data quality outcomes are directly comparable for adolescents and young adults, supporting the application of existing feasibility evidence for the Dreem3 across these age groups.

Finally, between-night stability did not significantly differ by age group. Most sleep outcomes demonstrated good-to-moderate stability. Consistent with gold standard PSG, sleep spindles and N3% were among the most stable features between nights (Levendowski et al., 2017). This result is promising and suggests that the Dreem3 may be comparable to PSG when it comes to accurately capturing more stable elements of sleep architecture. In a study examining sleep architecture in adolescents with PSG, ICC analyses found moderate to good within-subject stability across multiple recording nights (Ong et al., 2019). These findings align with our ICC analyses, demonstrating several trait-like characteristics of adolescent sleep. However, several sleep characteristics had poorer between-night stability (i.e., lower ICCs). We did not observe a difference in studies that involved at home sleep vs lab visit sleep **(Table S4).** Further research is needed to determine the number of nights required to obtain stability for all sleep architecture variables.

The present study is the first to evaluate age differences in portable sleep EEG device acceptability, feasibility, and data quality, however several limitations should be considered. First, our sample size and demographic characteristics may limit generalizability. Thirdly, we do not have sleep quality ratings from non-Dreem3 nights. Finally, missing UX data may have hindered our ability to discern between-group differences in acceptability. Future studies should test the Dreem3 in larger populations of young people and standardize recordings to either the lab or home environment. Additionally, more frequent and detailed assessments of user experience could inform product development and help future investigators explore how individuals adapt to the portable sleep EEG devices over repeated nights of wear.

These results support the acceptability, feasibility, and stability of Dreem3 recordings in both adolescent and young adult populations. While sources of sleep quality differences between age groups warrant further characterization, this pilot study suggests that portable sleep EEG devices may be a promising, scalable approach for capturing sleep physiology in adolescence and young adulthood. Future studies could apply these portable sleep recording technologies to understand neurodevelopmental variability in sleep physiology and how it contributes to health trajectories in larger, more generalizable developmental cohorts. Finally, we hope to conduct future studies validating the Dreem3 headband relative to gold-standard PSG in adolescents.

## Data Availability

Data produced in the present study can be shared upon reasonable request to the authors, with approval from the investigators on the contributing studies and appropriate regulatory bodies.

