## Supplemental Materials for "Acceptability, Feasibility, and Day-to-Day Stability of a Portable Sleep Recording Device in Adolescents versus Young Adults"

| **Table S1.** Participant demographic and clinical characteristics, N=81 | | | |
| --- | --- | --- | --- |
| **Variable** | **PITT– Study 1**  (n=21) | **PITT-Study 2**  (n=42) | **BCH**  (n=18) |
|  | *Mean(SD) or N(%)* | *Mean(SD) or N(%)* | *Mean(SD) or N(%)* |
| Age (yr) | 14.77 (1.03) | 21.93 (2.12) | 18.36 (6.18) |
| Female at Birth | 9 (42.86) | 21 (42.86) | 15 (83.33) |
| Hispanic/Latino | 3 (14.29) | 0 (0.00) | 2 (11.11) |
| Race |  |  |  |
| Asian | 0 (0.00) | 11 (25.58) | 5 (27.78) |
| Black | 2 (9.52) | 5 (11.63) | 0 (0.00) |
| Multiple Races | 1(4.76) | 3 (6.98) | 4 (22.22) |
| White | 18 (85.71) | 23 (53.49) | 9 (50.00) |
| Psychotropic Medication (y/n) | n | y | n |

*Note.* yr = year; y/n = yes/no; SD= Standard deviation; PITT = University of Pittsburgh; BCH = Boston Children’s Hospital; N = number

| **Table S2.** Pearson correlations between age and UX form ratings | | |
| --- | --- | --- |
| **UX Variable** | *r-value* | *p-value* |
| Mobile application was easy to use (Easy App) | 0.15 | 0.22 |
| Dreem3 was easy to set up in the mobile application (Easy to Setup) | -0.16 | 0.21 |
| Dreem3 was easy to sleep with on the first night (Easy to Sleep) | -0.08 | 0.50 |
| Dreem3 was comfortable (Comfortable) | -0.14 | 0.27 |
| Dreem3 was easy to put on (Easy to Put On) | -0.14 | 0.27 |
| Dreem3 was easy to use (Easy to Use) | -0.14 | 0.25 |
| Would recommend Dreem3 to a friend (Recommend) | -0.04 | 0.78 |
| Level of satisfaction wearing Dreem3 (Satisfaction) | 0.03 | 0.80 |
| Impact of Dreem3 on Sleep Quality | -0.28 | **0.02** |
| Impact of Dreem3 on Daytime Mood | -0.16 | 0.21 |
| Impact of Dreem3 on Daytime Alertness | 0.07 | 0.60 |
| Impact of Dreem3 on Day time Sleepiness | -0.16 | 0.20 |
| Impact of Dreem3 on Daytime Irritability | 0.04 | 0.77 |
| Impact of Dreem3 on Daytime Concentration | -0.17 | 0.18 |

*Note.* Dreem3 = Dreem Headband

| **Table S3:** Between-night stability (ICC) of sleep estimates by age group for participant with 2 nights of good quality sleep data F8-O1 (n=61). | | | | | |
| --- | --- | --- | --- | --- | --- |
| **Sleep Variable** | **Adolescents (A, n=24)** | | **Young Adults (B, n=37)** | | *Significant Difference* |
|  | *Mean ICC* | *95% CI* | *Mean ICC* | *95% CI* |  |
| N1% | 0.55 | (0.08-0.81) | 0.62 | (0.60-0.79) | ns |
| N2% | 0.57 | (0.09-0.80) | 0.47 | (0.26-0.72) | ns |
| N3% | 0.46 | (0.04-0.74) | 0.49 | (0.38-0.73) | ns |
| TST Hours | 0.27 | (0.01-0.69) | 0.26 | (0.00-0.29) | ns |
| TWT Hours | 0.27 | (0.01-0.65) | 0.26 | (0.11-0.54) | ns |
| REM% | 0.42 | (0.03-0.72) | 0.38 | (0.16-0.62) | ns |
| NREM Delta Rel Power | 0.63 | (0.25-0.83) | 0.55 | (0.52-0.75) | ns |
| NREM Theta Rel Power | 0.19 | (0.00-0.58) | 0.55 | (0.44-0.72) | ns |
| NREM Alpha Rel Power | 0.65 | (0.27-0.84) | 0.47 | (0.30-0.67) | ns |
| NREM Sigma Rel Power | 0.59 | (0.13-0.84) | 0.53 | (0.40-0.78) | ns |
| NREM Beta Rel Power | 0.40 | (0.01-0.69) | 0.20 | (0.02-0.47) | ns |
| NREM Delta Power | 0.34 | (0.02-0.63) | 0.56 | (0.55-0.76) | ns |
| NREM Theta Power | 0.26 | (0.00-0.63) | 0.40 | (0.25-0.65) | ns |
| NREM Alpha Power | 0.41 | (0.03-0.70) | 0.38 | (0.26-0.62) | ns |
| NREM Sigma Power | 0.44 | (0.03-0.72) | 0.33 | (0.33-0.55) | ns |
| NREM Beta Power | 0.22 | (0.00-0.57) | 0.23 | (0.06-0.47) | ns |
| REM Delta Rel Power | 0.28 | (0.00-0.72) | 0.41 | (0.26-0.63) | ns |
| REM Theta Rel Power | 0.17 | (0.00-0.58) | 0.37 | (0.23-0.64) | ns |
| REM Alpha Rel Power | 0.28 | (0.0-0.61) | 0.45 | (0.24-0.65) | ns |
| REM Sigma Rel Power | 0.46 | (0.10-0.71) | 0.37 | (0.21-0.65) | ns |
| REM Beta Rel Power | 0.35 | (0.08-0.69) | 0.43 | (0.23-0.69) | ns |
| REM Delta Power | 0.22 | (0.01-0.61) | 0.32 | (0.12-0.59) | ns |
| REM Theta Power | 0.29 | (0.01-0.65) | 0.40 | (0.17-0.65) | ns |
| REM Alpha Power | 0.35 | (0.11-0.61) | 0.46 | (0.23-0.68) | ns |
| REM Sigma Power | 0.34 | (0.07-0.62) | 0.33 | (0.10-0.58) | ns |
| REM Beta Power | 0.27 | (0.06-0.60) | 0.18 | (0.02-0.46) | ns |
| Spindle Amplitude (Slow) | 0.51 | (0.10-0.79) | 0.39 | (0.32-0.63) | ns |
| Spindle Density (Slow) | 0.63 | (0.20-0.83) | 0.53 | (0.44-0.75) | ns |
| Spindle Duration (Slow) | 0.59 | (0.14-0.79) | 0.35 | (0.12-0.60) | ns |
| Integrated Spindle Activity (Slow) | 0.67 | (0.16-0.86) | 0.48 | (0.42-0.68) | ns |
| CHIRP (Slow) | 0.72 | (0.37-0.90) | 0.48 | (0.35-0.70) | ns |
| Fast Fourier Transform (Slow) | 0.78 | (0.47-0.92) | 0.60 | (0.50-0.80) | ns |
| Spindle Amplitude (Fast) | 0.42 | (0.07-0.75) | 0.24 | (0.07-0.52) | ns |
| Spindle Density (Fast) | 0.50 | (0.06-0.74) | 0.66 | (0.78-0.81) | ns |
| Spindle Duration (Fast) | 0.23 | (0.00-0.55) | 0.18 | (0.00-0.48) | ns |
| Integrated Spindle Activity (Fast) | 0.42 | (0.05-0.69) | 0.48 | (0.43-0.68) | ns |
| CHIRP (Fast) | 0.16 | (0.04-0.49) | 0.49 | (0.37-0.72) | ns |
| Fast Fourier Transform (Fast) | 0.72 | (0.39-0.88) | 0.46 | (0.19-0.68) | ns |
| SO Duration | 0.44 | (0.06-0.76) | 0.48 | (0.40-0.70) | ns |
| SO Peak-to-Peak | 0.29 | (0.0-0.59) | 0.51 | (0.44-0.69) | ns |
| SO Rate | 0.46 | (0.10-0.72) | 0.30 | (0.06-0.56) | ns |
| SO Negative Slope | 0.23 | (0.00-0.60) | 0.53 | (0.41-0.72) | ns |

Note. ICC = Intraclass Correlation Coefficient; REM = Rapid Eye Movement; NREM = Non-Rapid Eye Movement; N1 = NREM Stage 1; N2 = NREM Stage 2 ;N3 = NREM Stage 3; TST = Total Sleep Time; TWT = Total Wake Time; Rel= Relative; SO = Slow Oscillation ; CHIRP = spindle frequency modulation; bootstrapped confidence intervals based on 500 iterations.

| **Table S4.** UX survey acceptability data by age group adjusted for race, weekday/weekend, and psychotropic medication use, N=66 | | | | | | |
| --- | --- | --- | --- | --- | --- | --- |
| **UX Variable** | **Adolescents**  **(9-17y; n=24)** | | **Young Adults**  **(18-26y; n=42)** | | **Statistic** | |
|  | *Mean* | *SD* | *Mean* | *SD* | *F-statistic* | *p-value* |
| Mobile application was easy to use (Easy App)^a^ | 6.25 | 1.03 | 6.60 | 1.01 | 1.36 | 0.18 |
| Dreem3 was easy to set up in the mobile application (Easy to Set Up)^a^ | 5.38 | 1.38 | 4.86 | 1.82 | -2.24 | **0.03** |
| Dreem3 was easy to sleep with on the first night (Easy to Sleep)^a^ | 5.13 | 1.57 | 4.76 | 1.39 | -1.14 | 0.26 |
| Dreem3 was comfortable (Comfortable)^a^ | 4.83 | 1.66 | 4.38 | 1.34 | -1.13 | 0.26 |
| Dreem3 was easy to put on (Easy to Put On)^a^ | 5.63 | 1.47 | 5.12 | 1.55 | -1.08 | 0.29 |
| Dreem3 was easy to use (Easy to Use)^a^ | 5.46 | 1.25 | 4.95 | 1.53 | -1.55 | 0.13 |
| Would recommend Dreem3 to a friend (Recommend)^a^ | 4.50 | 1.96 | 4.22 | 1.67 | -1.12 | 0.27 |
| Satisfaction wearing Dreem3 (Satisfaction)^a^ | 4.75 | 2.11 | 4.73 | 1.27 | -0.26 | 0.80 |
| Sleep Quality^b^ | -0.04 | 0.62 | -0.43 | 0.55 | -1.91 | *0.06* |
| Mood^b^ | 0.04 | 0.21 | 0.00 | 0.22 | 0.11 | 0.91 |
| Alertness^b^ | 0.00 | 0.30 | 0.07 | 0.34 | 1.59 | 0.12 |
| Sleepiness^b^ | 0.13 | 0.34 | 0.02 | 0.35 | -0.50 | 0.62 |
| Irritability^b^ | 0.00 | 0.30 | 0.05 | 0.31 | 0.76 | 0.45 |
| Concentration^b^ | 0.17 | 0.49 | 0.05 | 0.31 | -0.76 | 0.45 |

*Note.* For the first eight items, scales ranged from 1 - 7. For the last six items, scales ranged from –2 – 2.

**Figure S1**

*Dreem UX Survey*

**Dreem Satisfaction Questionnaire**

The following questionnaire is part of our evaluation of the Dreem3 sleep recording headband that you used to track your sleep. We encourage you to answer each question as honestly as possible so that we may accurately evaluate your current level of satisfaction with your experience.

**Please answer the following questions so we can learn a little more about you:**

**Age:** ________ (years)

**Sex at birth:** M	F

**Ethnicity:** Hispanic/Latino	Not Hispanic/Latino

**Race:** White	Black	Asian	Native American/Pacific Islander	Multiple	Other

**Typically, my sleep quality is:**

| 1 | 2 | 3 | 4 | 5 | 6 | 7 |
| --- | --- | --- | --- | --- | --- | --- |
| Very poor |  |  | okay |  |  | Very good |

**For each question below, please circle the response that best describes how you honestly feel:**

How easy was it download the *Alfin* app?

| 1 | 2 | 3 | 4 | 5 | 6 | 7 |
| --- | --- | --- | --- | --- | --- | --- |
| Not at all |  |  | somewhat |  |  | Very much |

How easy was it to set up the Dreem3 headband in the *Alfin* app?

| 1 | 2 | 3 | 4 | 5 | 6 | 7 |
| --- | --- | --- | --- | --- | --- | --- |
| Not at all |  |  | somewhat |  |  | Very much |

How easy was it to sleep with the Dreem3 headband on the first night?

| 1 | 2 | 3 | 4 | 5 | 6 | 7 |
| --- | --- | --- | --- | --- | --- | --- |
| Not at all |  |  | somewhat |  |  | Very much |

Was it easier to sleep with the Dreem3 headband **after** the first night?

| 1 | 2 | 3 | 4 | 5 | 6 | 7 |
| --- | --- | --- | --- | --- | --- | --- |
| Not at all |  |  | somewhat |  |  | Very much |

How comfortable was the Dreem3 headband?

| 1 | 2 | 3 | 4 | 5 | 6 | 7 |
| --- | --- | --- | --- | --- | --- | --- |
| Not at all |  |  | somewhat |  |  | Very much |

How easy was it to put on the Dreem3 headband?

| 1 | 2 | 3 | 4 | 5 | 6 | 7 |
| --- | --- | --- | --- | --- | --- | --- |
| Not at all |  |  | somewhat |  |  | Very much |

How easy was it to use the Dreem3 headband?

| 1 | 2 | 3 | 4 | 5 | 6 | 7 |
| --- | --- | --- | --- | --- | --- | --- |
| Not at all |  |  | somewhat |  |  | Very much |

The 3 nights of sleep tracking with the Dreem3 headband was:

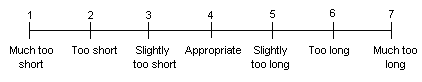

Would you recommend the Dreem3 sleep recording headband to a friend?

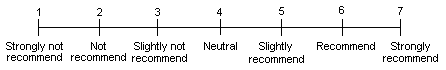

Overall, my level of satisfaction with my experience wearing the Dreem3 headband is:

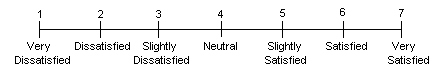

Please rate how wearing the Dreem3 headband impacted your….

|  | Much worse | A little worse | No change | A little better | Much better |
| --- | --- | --- | --- | --- | --- |
| Sleep Quality | -2 | -1 | 0 | 1 | 2 |
| Mood | -2 | -1 | 0 | 1 | 2 |
| Alertness | -2 | -1 | 0 | 1 | 2 |
| Sleepiness during the day / falling asleep | -2 | -1 | 0 | 1 | 2 |
| Getting along with others / irritability | -2 | -1 | 0 | 1 | 2 |
| Ability to concentrate | -2 | -1 | 0 | 1 | 2 |

10. In your opinion, what can we do to improve the experience of wearing the Dreem3 sleep recording headband?

Please let us know any other feedback you have about your experience wearing the Dreem3 sleep recording headband:

11. We welcome any additional comments. Thank you!!

**Figure S2**

*Recording quality by participant, group, and night*

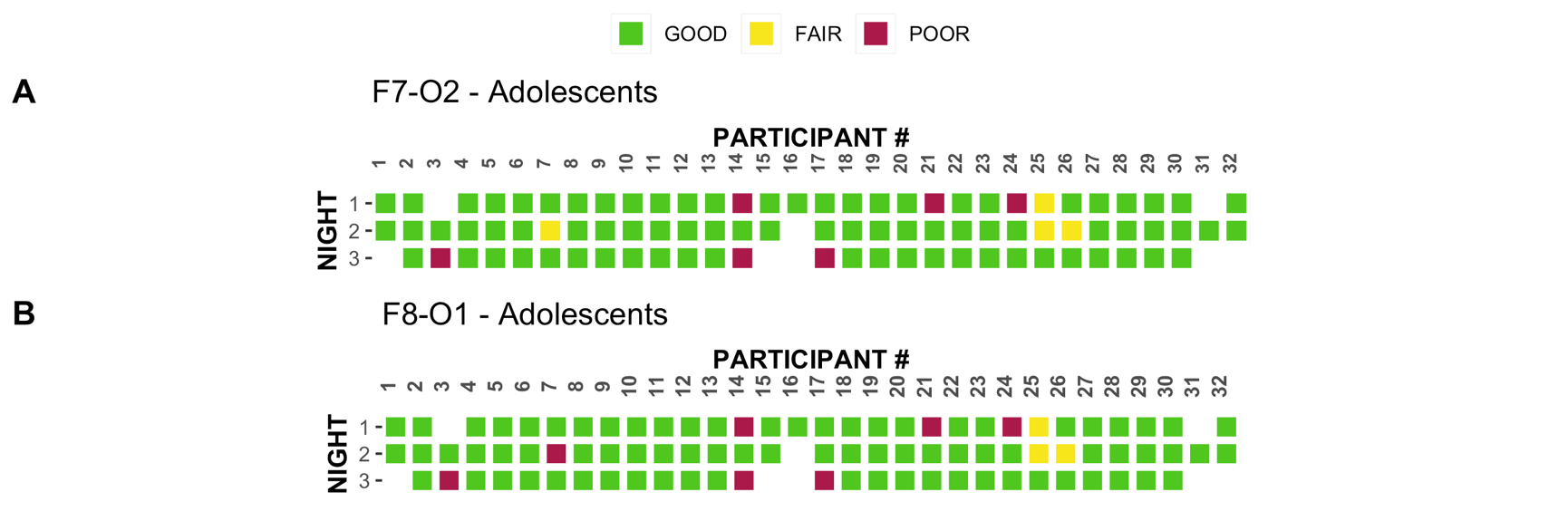

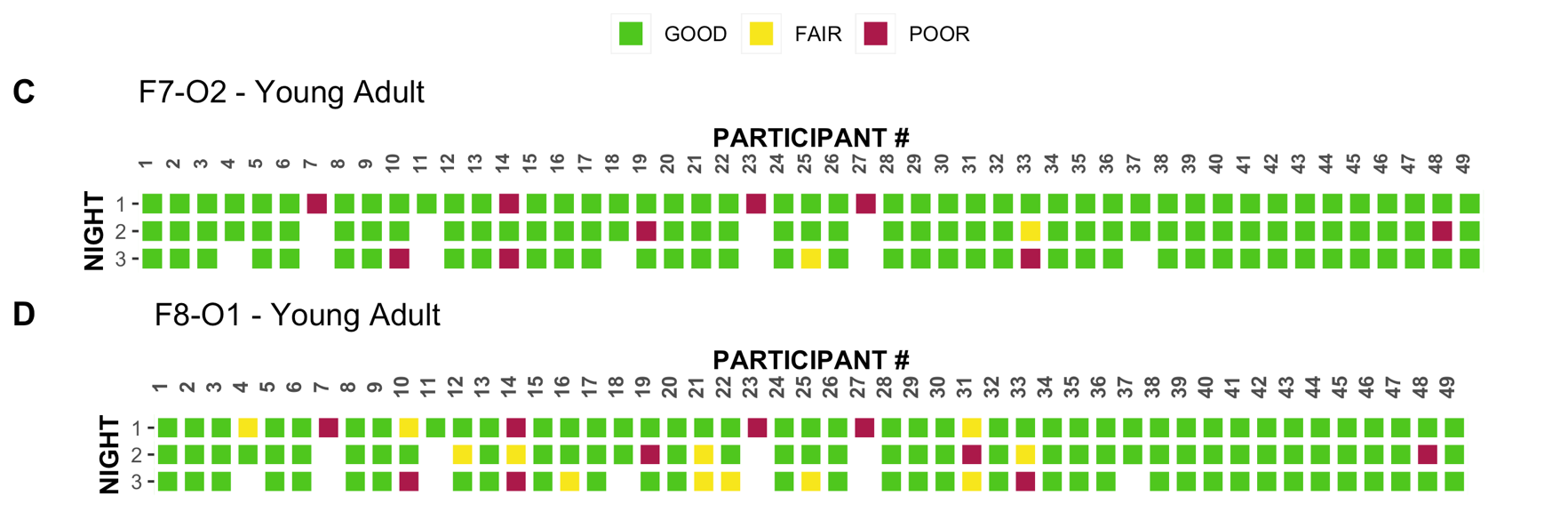

*Note.* Panels A and B represent signal quality among adolescents for electrodes F7-O2 and F8-O1. Panels C and D represent signal quality among young adults for electrodes F7-O2 and F8-O1.

**Figure S3**

*Between-night stability (ICC) of sleep estimates by age group for participants with at least 3 nights of good quality sleep data (F7-O2 & F8-O1 derivations)*

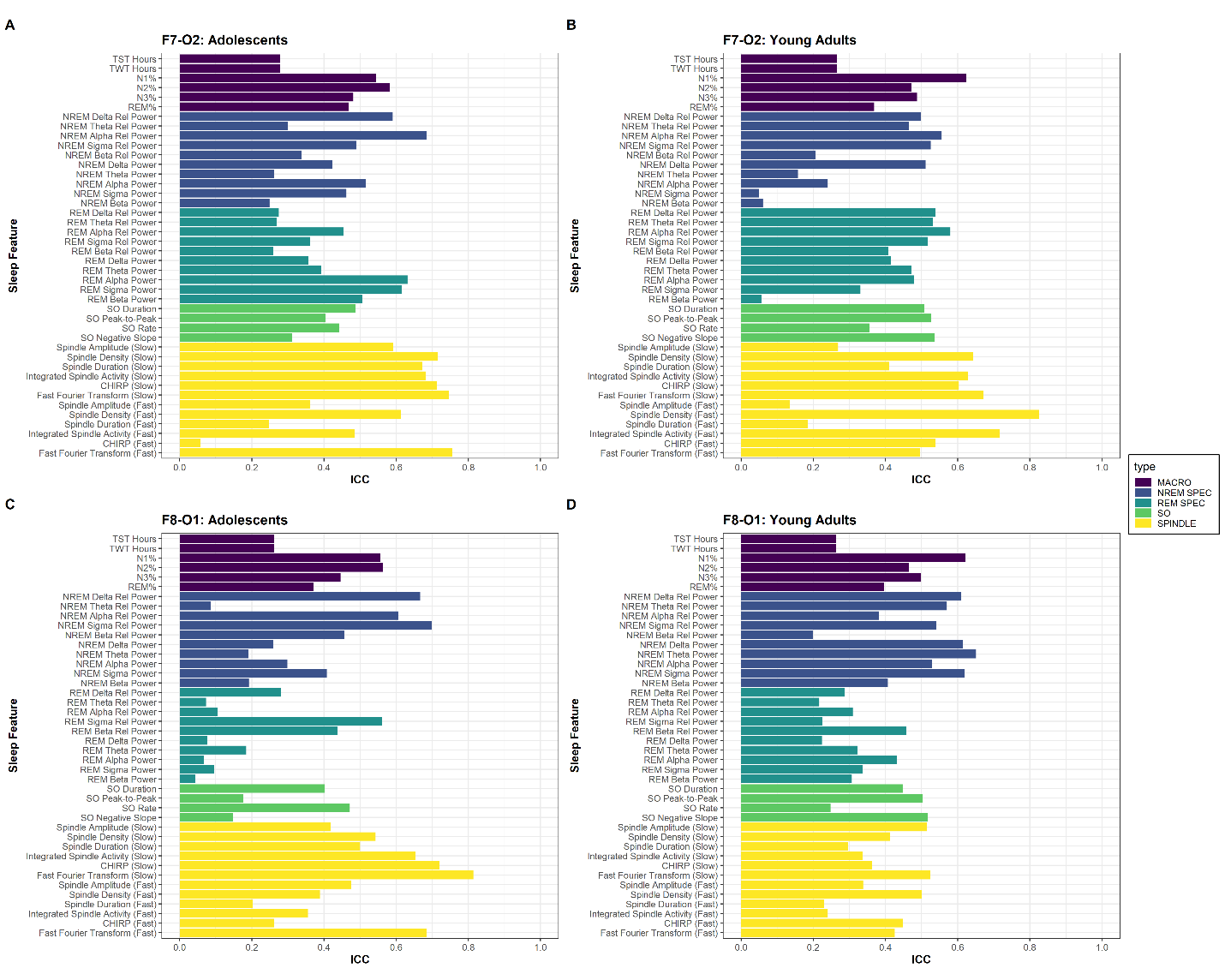
*Note.* F7-O2 and F8-O1 ICC plots combined for Adolescents and Young Adults. **A:** F7-O2 Adolescents (n=29). **B:** F7-O2 Adults (n=43). **C:** F8-O1 Adolescents (n=24). **D:** F8-O1 Adults(n=37). ICC = Intraclass Correlation Coefficient; REM = Rapid Eye Movement; NREM = Non-Rapid Eye Movement; N1 = NREM Stage 1; N2 = NREM Stage 2 ;N3 = NREM Stage 3; TST = Total Sleep Time; TWT = Total Wake Time; Rel= Relative; SO = Slow Oscillation ; CHIRP = spindle frequency modulation; bootstrapped confidence intervals based on 500 iterations.

**Table S4**

| **Table S4:** Between-night stability (ICC) of sleep estimates by PSG for participants with at least 2 nights of good quality sleep data for F7-O2 (n=72). | | | | | |
| --- | --- | --- | --- | --- | --- |
| **Sleep Variable** | **PSG (A, n=36)** | | **No PSG (B, n=36)** | | *Significant Difference* |
|  | *Mean ICC* | *95% CI^a^* | *Mean ICC* | *95% CI^a^* |  |
| N1% | 0.60 | (0.03-0.56) | 0.64 | (0.29-0.80) | ns |
| N2% | 0.47 | (0.13-0.80) | 0.48 | (0.13-0.20) | ns |
| N3% | 0.47 | (0.17-0.70) | 0.47 | (0.17-0.70) | ns |
| REM% | 0.35 | (0.09-0.57) | 0.35 | (0.09-0.57) | ns |
| TST Hours | 0.31 | (0.05-0.54) | 0.31 | (0.05-0.50) | ns |
| TWT Hours | 0.31 | (0.03-0.56) | 0.31 | (0.03-0.56) | ns |
| NREM Delta Rel Power | 0.53 | (0.23-0.74) | 0.53 | (0.23-0.75) | ns |
| NREM Theta Rel Power | 0.45 | (0.16-0.67) | 0.49 | (0.16-0.67) | ns |
| NREM Alpha Rel Power | 0.58 | (0.25-0.76) | 0.58 | (0.25-0.76) | ns |
| NREM Sigma Rel Power | 0.54 | (0.24-0.78) | 0.54 | (0.24-0.78) | ns |
| NREM Beta Rel Power | 0.23 | (0.00-0.41) | 0.23 | (0.00-0.52) | ns |
| NREM Delta Power | 0.62 | (0.33-0.78) | 0.61 | (0.33-0.78) | ns |
| NREM Theta Power | 0.15 | (0.00-0.41) | 0.15 | (0.00-0.41) | ns |
| NREM Alpha Power | 0.23 | (0.01-0.49) | 0.23 | (0.01-0.49) | ns |
| NREM Sigma Power | 0.05 | (0.00-0.31) | 0.05 | (0.00-0.49) | ns |
| NREM Beta Power | 0.06 | (0.00-0.27) | 0.06 | (0.00-0.27) | ns |
| REM Delta Rel Power | 0.54 | (0.04-0.75) | 0.54 | (0.04-0.75) | ns |
| REM Theta Rel Power | 0.53 | (0.23-0.72) | 0.53 | (0.23-0.72) | ns |
| REM Alpha Rel Power | 0.55 | (0.27-0.75) | 0.55 | (0.27-0.75) | ns |
| REM Sigma Rel Power | 0.52 | (0.21-0.72) | 0.52 | (0.21-0.72) | ns |
| REM Beta Rel Power | 0.40 | (0.05-0.62) | 0.40 | (0.05-0.60) | ns |
| REM Delta Power | 0.38 | (0.11-0.59) | 0.38 | (0.11-0.59) | ns |
| REM Theta Power | 0.41 | (0.10-0.69) | 0.45 | (0.10-0.69) | ns |
| REM Alpha Power | 0.42 | (0.17-0.71) | 0.46 | (0.17-0.71) | ns |
| REM Sigma Power | 0.32 | (0.00-0.58) | 0.32 | (0.00-0.58) | ns |
| REM Beta Power | 0.05 | (0.00-0.37) | 0.05 | (0.00-0.37) | ns |
| Spindle Amplitude (Slow) | 0.26 | (0.01-0.49) | 0.26 | (0.02-0.49) | ns |
| Spindle Density (Slow) | 0.63 | (0.11-0.80) | 0.63 | (0.11-0.80) | ns |
| Spindle Duration (Slow) | 0.41 | (0.09-0.64) | 0.41 | (0.09-0.64) | ns |
| Integrated Spindle Activity (Slow) | 0.60 | (0.23-0.77) | 0.60 | (0.23-0.77) | ns |
| CHIRP (Slow) | 0.57 | (0.26-0.79) | 0.57 | (0.26-0.79) | ns |
| Fast Fourier Transform (Slow) | 0.65 | (0.28-0.80) | 0.65 | (0.28-0.80) | ns |
| Spindle Amplitude (Fast) | 0.12 | (0.00-0.38) | 0.12 | (0.00-0.38) | ns |
| Spindle Density (Fast) | 0.81 | (0.51-0.91) | 0.81 | (0.51-0.91) | ns |
| Spindle Duration (Fast) | 0.19 | (0.00-0.45) | 0.19 | (0.00-0.45) | ns |
| Integrated Spindle Activity (Fast) | 0.70 | (0.33-0.86) | 0.70 | (0.33-0.86) | ns |
| CHIRP (Fast) | 0.51 | (0.17-0.70) | 0.51 | (0.17-0.70) | ns |
| Fast Fourier Transform (Fast) | 0.44 | (0.19-0.68) | 0.44 | (0.19-0.68) | ns |
| SO Duration | 0.53 | (0.22-0.73) | 0.53 | (0.22-0.73) | ns |
| SO Peak-to-Peak | 0.55 | (0.22-0.72) | 0.55 | (0.22-0.72) | ns |
| SO Rate | 0.37 | (0.06-0.66) | 0.37 | (0.06-0.66) | ns |
| SO Negative Slope | 0.58 | (0.00-0.73) | 0.65 | (0.33-0.82) | ns |

Note. ICC = Intraclass Correlation Coefficient; REM = Rapid Eye Movement; NREM = Non-Rapid Eye Movement; N1 = NREM Stage 1; N2 = NREM Stage 2 ;N3 = NREM Stage 3; TST = Total Sleep Time; TWT = Total Wake Time; Rel= Relative; SO = Slow Oscillation ; CHIRP = spindle frequency modulation; bootstrapped confidence intervals based on 500 iterations.
